# Data Veracity of Patients and Health Consumers Reported Adverse Drug Reactions on Twitter: Key Linguistic Features, Twitter Variables, and Association Rules

**DOI:** 10.1101/2020.11.03.20225532

**Authors:** Tianchu Lyu, Andrew Eidson, Jungmi Jun, Xiajie Zhou, Xiang Cui, Chen Liang

## Abstract

Adverse drug reactions (ADRs) lead to high disease burden and health expenditure. Aside from traditional data sources used for pharmacovigilance, social media have emerged as an important supplemental data source for monitoring patients and consumers reported ADRs. Recently, there have been increasing concerns about the data veracity of ADRs extracted from social media. Our objective is to categorize different levels of data veracity and explore influential linguistic features and Twitter variables as they may be used for screening ADRs for high data veracity. We annotated a corpus of ADRs with linguistic features validated by clinical experts. Multinomial logistic regression was applied to investigate the associations between the linguistic features and levels of data veracity. We found that using first-person pronouns, expressing negative sentiment, ADR and drug name being in the same sentence were significantly associated with higher levels of data veracity (all p < 0.05), using medical terminology and less indications were associated with good data veracity (p < 0.05), less drug numbers were marginally associated with good data veracity (p = 0.053). These findings suggest an opportunity of developing machine learning models for automatic screening of ADRs from Twitter using identified key linguistic features, Twitter variables, and association rules.

## Introduction

Adverse drug reactions (ADRs) have been one of the leading causes of morbidity and mortality in the United States and a significant cost driver, accounting for about 30.1 billion US dollars of annual health expenditure.^1,2^ The World Health Organization (WHO) defined the ADR as “a response to a drug which is noxious and unintended, and which occurs at doses normally used in man for the prophylaxis, diagnosis, or therapy of disease, or for the modifications of physiological function”.^3^ According to the report from the Institute of Medicine, ADRs account for 7,000 deaths among 44,000 to 98,000 deaths caused by medical errors annually.^4^ The annual rates of ADR-related deaths have increased from 1999 to 2006, ranging from 8 to 12 per 1,000 people.^5^ In addition, 0.5% to 12.04% of hospital admissions were associated with ADRs, with higher rates in elderly patients.^6-8^ Age and the number of medication were identified as risk factors for ADR in outpatients,^9^ suggesting that the cost concentration issue in health expenditure was aggravated by ADRs since elderly people are more likely to receive various medications because of chronic diseases.^10^

The importance in monitoring ADRs and other drug-related problems, denoted as pharmacovigilance (PV), was recognized for the first time during the thalidomide disaster in 1961. Since then, PV has been widely used to detect, assess, and understand ADRs.^11^ Over the decades, health care providers and stakeholders have been seeking ways to improve PV and have identified problems limited by the data sources of PV. Traditional sources of data used for identifying ADRs include clinical trials, pharmaceutical industry reports, and spontaneous reporting systems (SRSs).^12-14^ Electronic health records (EHR) have also been used as a promising data source for PV in recent years.^15,16^ However, these data sources may be limited in terms of capturing the full spectrum of ADRs along the course of health care and social care. For example, clinical trials may fail to detect rare ADRs because of their inherent shortcomings including relatively short duration and small sample sizes.^17^ SRSs include information only from standardized reports and, therefore, suffer from under-reporting problems.^18^ Additionally, there are barriers (e.g., poor coordination and privacy concerns) for organizations to integrate health data from diverse health information systems across different health care interest groups and to exchange health information among the groups.^19,20^ It is also particularly challenging to acquire ADR data from hard-to-reach patients and health consumers who have experienced ADRs but have not interacted with any sector of health care system.^1,21^

Social media have emerged as a viable data source for collecting ADRs reported by patients and consumers and an important supplement for traditional PV because they contain rich patients and consumers-provided information.^22^ Avery and colleagues suggested that types and adverse effects of drugs reported by patients are different from that reported by health care professionals and patient-reported data contain more details.^23^ In comparison to the traditional data sources, social media platforms, such as Twitter, are much easier and less costly to access, and produce richer data abundance.^24^ Recent studies have demonstrated the feasibility of using social media as a promising data source for post-marketing PV. It was reported that social media data can be utilized for validating pre-existing ADRs as well as detecting signals of new or rare ADRs.^25^ Abou Taam et al. examined ‘benfluorex’ related content on three social media websites and found various ADRs such as anxiety, anger, and valvulopathy.^26^ Yang et al.’s analysis of social media content (e.g., MedHelp, PatientsLikeMe) and identified drug-ADR pairs, such as ‘Lansoprazole’ and ‘diarrhea’, ‘Prozac’ and ‘depression’, and ‘Luvox’ and ‘heart disease’.^27^ Pierce et al. examined ten safety signals and found that ‘dronedarone–vasculitis’ exhibited in social media prior to FDA signal detection, suggesting that social media listening could contribute to early detection of ADRs.^28^

Despite these advantages of using social media data for PV, uneven data veracity has been a major concern. Researchers have explored the data veracity issue from diverse perspectives. Hoang and colleagues measured data uncertainty and rarity for ADRs extracted from Twitter, and proposed authenticity and credibility as two root causes of poor data veracity.^29,30^ Nguyen et al. evaluated trustworthiness to improve the accuracy of social media data.^31^ In addition, data veracity problems were reported to threaten availability, confidentiality, and integrity of social media data and analyses.^32^ Building on the existing literature, we summarized three obstacles to good data veracity on social media when it comes to the identification of ADRs. First, most patients and health consumers are laypersons who tend to use their own terms in describing a health issue on social media, which often differ from terminologies that are validated and used by healthcare professionals such as Unified Medical Language System (UMLS).^33^ Second, social media users may not constantly provide credible information as regulatory individual case safety reports (ICSR) provided by pharmaceutical and biotechnology companies.^34^ Third, due to the lack of medical knowledge, laypersons may not be able to correctly link an ADR to the corresponding medication. Oftentimes, they have difficulties in distinguishing ADRs from comorbidities or indications.^30^ Collectively, these obstacles contribute to the uneven data veracity of consumer reported ADRs on social media, which significantly impede novel PV research using social media data. The process of identifying levels of data veracity is labor intensive because the task requires dissecting improvised social media posts in free text, often complicated by frequent typos, copy-forwarding, and context-dependent language.

Recent advances in data science have introduced natural language processing (NLP) methods to overcome the challenges of analyzing written communications on social media. NLP has provided a feasible way to clean and prepare the textual social media data before being used for PV, including spelling correction, lemmatization, lowercasing, annotation, etc..^35^ Using linguistic features such as first-person pronoun (i.e., “I”, “me”, “my”, etc.) or URL could help improve accuracy of extracting ADR messages.^36,37^ NLP also showed potentials to recognize, extract, and quantify subjective experiences from Twitter users, which is often denoted as sentiment analysis.^37,38^ Despite the fast development of NLP approaches used for identifying ADR-related tweets, linguistic features and methods tailored for data veracity of ADRs on social media are sparse.

While there has been increasing attention devoted to the data veracity issue of ADR-related social media,^29,39^ no existing studies have systematically defined and characterized data veracity of ADRs on social media in the context of using social media data for PV. In this study, we propose a viable approach to identify different levels of data veracity for ADRs extracted from Twitter and compared levels of data veracity with different linguistic features and Twitter variables (e.g., number of followers, retweets, user-reported demographics). To this end, we integrated existing Twitter variables and nuanced linguistic features extracted from Twitter posts to establish multinomial logistic regression models where we aimed to identify factors (i.e., Twitter variables and linguistic features) that contribute most to specific levels of ADR data veracity. Twitter variables are extracted through Twitter API. Linguistic features of Twitter posts are extracted by employing an annotation protocol that is clinically plausible for screening ADR events with different veracity levels. Anticipated outcomes of this study hold promise to inform the future development of a machine learning model that can automatically screen high-veracity ADRs detected from Twitter, which will greatly facilitate PV research using social media data.

## Materials and Methods

### Data Source

The data used for analysis were from a publicly available corpus of tweets containing ADRs, indexed by tweeter IDs.^36^ The corpus consists of 10,822 tweets that were annotated with medications and indications. There are 1,217 tweets containing at least one ADR. We retrieved all the available ADR tweets by twitter IDs via the Twitter API implemented with Python in June of 2020. 766 of tweets were no longer retrievable since the corpus was created in 2014 and some tweets or user profiles had been deleted by the time we collected the data. As a result, we retrieved 451 tweets labeled in the corpus as “containing an ADR” for analyses. For validating drug-ADR pairs in the corpus, we used SIDER 4.1 database since it is widely recognized in PV research.^40^ Drug-ADR pairs identified in the corpus were compared against the clinically validated drug-ADR pairs documented in SIDER 4.1.

### Annotation

We performed an annotation task to identify levels of data veracity as well as ten critical linguistic features from the tweets. A panel of two clinical experts (AE and TL) was assembled to perform the annotation. The two clinical experts have received proper training in pharmacology and ADR-related work from medical schools. We defined an ADR as “an undesired effect of the drug experienced by the patient” and an indication as “the sign, symptom, syndrome, or disease that is the reason or the purpose for the patient taking the drug or is the desired primary effect of the drug”.^35^ Based on the key linguistic features used in prior studies for identifying ADRs on Twitter, the expert panel developed a protocol for the annotation (Figure 1). Each panel expert followed this protocol and independently performed the annotation task. The interrater reliability was measured by Cohen’s Kappa Statistic using data veracity level as representative measurement. The calculated Kappa value is 0.80, which is considered high agreement.^41^ Disagreement in annotations was resolved during the panel discussion. Below we summarized the key components of the protocol including data veracity levels and key linguistic features.

**Figure 1.**
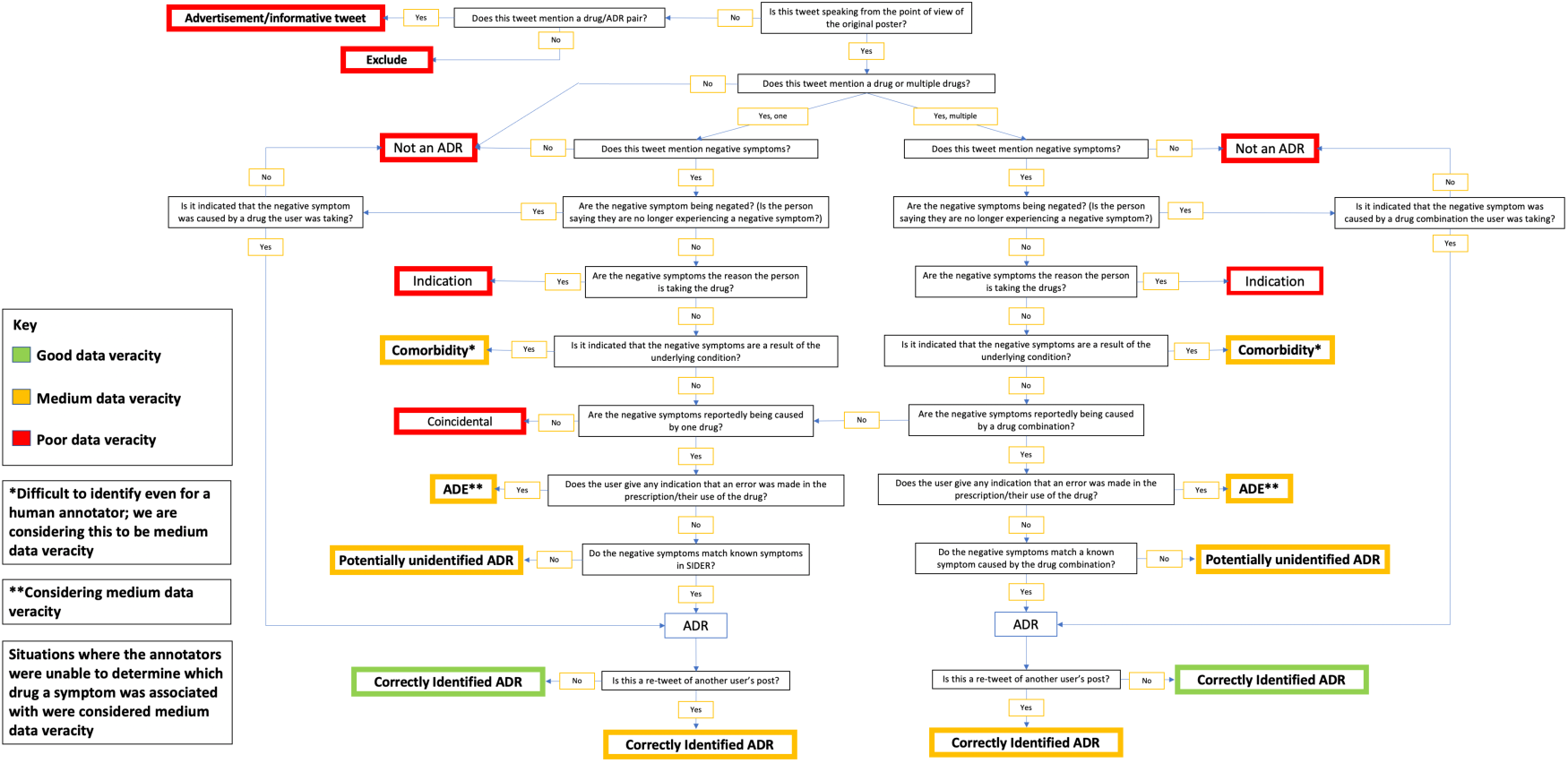
Annotation flow chart.

#### Data Veracity Levels

Data veracity of a tweet is defined in three levels including poor, moderate, and good data veracity. To be annotated as good data veracity, the tweets should (1) be correctly identified as an ADR tweet in the corpus, (2) not be a retweet, (3) contain no URL, (4) be first-person experience, (5) explicitly state the drug name and corresponding ADRs, and (6) correctly include the indications if any. A tweet was classified as moderate data veracity when the tweet did not contain sufficient information required for determining good data veracity. Specifically, moderate data veracity was identified when the expert was unable to decide whether the Twitter user reported an ADR they had experienced, or a single drug-ADR pair could not be identified. For example, tweets with moderate data veracity might contain a potential ADR that has been identified but was not listed in SIDER database. Tweets with poor data veracity could be expert views, advertisements, case reports, and tweets that contain URLs or indications that were mistakenly identified as ADRs. Table 1 is an operational classification system for data veracity generated based on the annotation protocol (Figure 1).

**Table 1.** Operational classification system for data veracity.

|  |  |
| --- | --- |
| Good data veracity | • Non-retweet, correctly identified ADR<br>• First-person experience<br>• Explicitly state the drug name and ADR<br>• No URLs<br>• Correctly identified indications (they are sometimes in the same tweet as an ADR) |
| Moderate data veracity | • Unable to determine if the users are reporting an ADR they have experienced<br>• Tweet referring to ADRs experienced by others<br>• Vague ADR descriptions that are unable to be paired with a particular ADR in SIDER<br>• Retweet, correctly identified ADR<br>• Not first-person experience<br>• Explicitly state the drug name and ADR word<br>• ADE identified as ADR<br>• Difficult to pick out sometimes, even for a medical expert<br>• Comorbidity identified as ADR<br>• Difficult to pick out sometimes, even for a medical expert<br>• Potential ADR identified that is not listed in SIDER<br>• The ADR is from two or more drugs working together<br>• Drug combination<br>• Took a different drug recently |
| Poor data veracity | • Expert views<br>• Listing ADRs of a drug instead of stating that they are experiencing them<br>• Contain URLs<br>• Advertisements<br>• Case reports<br>• Indications that were mis-identified as ADR |

#### Key Linguistic Features

- First-person, second-person, and third-person pronouns were annotated since they are indicative variables found in previous studies used for improving the accuracy of identification.^29,36,38^
- Sentiments were also annotated since prior studies found that most ADR tweets are associated with negative sentiment, and that sentiment analysis is commonly used to identify ADR-related tweets.^29,36-38,42^
- We annotated whether drug names and mentions of ADRs were in the same sentence.
- We annotated the presences of ADRs that are captured by medical terminology systems used by SIDER including Medical Dictionary for Regulatory Activities (MedDRA).
- We annotated the number of drugs and the number of indications.

### Multinomial Logistic Regression

Multinomial logistic regression was applied to explore the effects of different indicative and contra-indicative factors of interest on data veracity. We removed two variables before model selection: URL and retweet. These two variables were dropped because the extreme imbalance in the frequencies of values can lead to a quasi-complete separation of data. Specifically, there were only three tweets containing URLs and ten retweets. We used backwards selection method to select the final model. The likelihood ratio test was applied to compare the goodness of fit of the full model and the reduced models. Alternatively, we calculated the Akaike Information Criterion (AIC) to test the goodness of fit of the model. No obvious multicollinearity was observed, assessed using variance inflation factors (VIF), tolerance statistics, and eigenvalue and condition index. All the statistical analyses were performed using SAS 9.4.

## Results

Among the 451 tweets, 36 (7.98%), 196 (43.46%), and 219 (48.56%) tweets are in poor, moderate, and good data veracity, correspondingly. Compared with tweets of good and moderate data veracity, poor data veracity tweets were less likely to use first-person pronoun (79.91%, 77.55%, and 41.67%, respectively), more likely to use second-person pronoun (6.85%, 15.31%, and 16.67%), less likely to present drug and ADR in the same sentence (71.69%, 81.63%, and 44.44%), and less likely to use medical terminology for ADRs (34.70%, 14.80%, and 13.89%). Poor data veracity tweets contain more indications. Specifically, the percentage of tweets in poor data veracity that contain one or more indications was 19.44%, compared with 6.12% for tweets in moderate data veracity and 4.57% for tweets in good data veracity. The percentage of tweets in poor data veracity that have no drug name mentioned was 16.67%, much higher than that of tweets in moderate and good data veracity (3.96% and 0.00%, respectively). Besides, tweets in poor data veracity were more likely to include more than one drug name than tweets in moderate and good data veracity (36.11%, 17.33%, and 13.24%). Equally, the percentage of tweets in poor data veracity containing only one drug name is the lowest across the three data veracity levels (47.22%, 78.71%, and 86.30%) (Table 2).

**Table 2.** Tweets characteristics by data veracity levels.

|  | Data Veracity Level |  |  |  |  |  | Total |
| --- | --- | --- | --- | --- | --- | --- | --- |
|  | Poor |  | Moderate |  | Good |  |  |
| Whether user uses first-person pronoun |  |  |  |  |  |  |  |
| Yes | 15 | (41.67%) | 152 | (77.55%) | 175 | (79.91%) | 342 |
| No | 21 | (58.33%) | 44 | (22.45%) | 44 | (20.09%) | 109 |
| Whether user uses second-person pronoun |  |  |  |  |  |  |  |
| Yes | 6 | (16.67%) | 30 | (15.31%) | 15 | (6.85%) | 51 |
| No | 30 | (83.33%) | 166 | (84.69%) | 204 | (93.15%) | 400 |
| Negative sentiment of ADR sentence |  |  |  |  |  |  |  |
| Yes | 7 | (19.44%) | 114 | (58.16%) | 132 | (60.27%) | 253 |
| No | 29 | (80.56%) | 82 | (41.84%) | 87 | (39.73%) | 198 |
| Whether drug and ADR are in the same sentence |  |  |  |  |  |  |  |
| Yes | 16 | (44.44%) | 160 | (81.63%) | 157 | (71.69%) | 333 |
| No | 20 | (55.56%) | 36 | (18.37%) | 62 | (28.31%) | 118 |
| Whether user uses medical terminology |  |  |  |  |  |  |  |
| Yes | 5 | (13.89%) | 29 | (14.80%) | 76 | (34.70%) | 110 |
| No | 31 | (86.11%) | 167 | (85.20%) | 143 | (65.30%) | 341 |
| Number of drugs |  |  |  |  |  |  |  |
| 0 | 6 | (16.67%) | 8 | (3.96%) | 0 | (0.00%) | 14 |
| 1 | 17 | (47.22%) | 159 | (78.71%) | 189 | (86.30%) | 365 |
| 2 | 12 | (33.33%) | 27 | (13.37%) | 27 | (12.33%) | 66 |
| 3 | 1 | (2.78%) | 8 | (3.96%) | 2 | (0.91%) | 11 |
| 4 | 0 | (0.00%) | 0 | (0.00%) | 1 | (0.46%) | 1 |
| Number of indications |  |  |  |  |  |  |  |
| 0 | 29 | (80.56%) | 184 | (93.88%) | 209 | (95.43%) | 422 |
| 1 | 7 | (19.44%) | 9 | (4.59%) | 9 | (4.11%) | 25 |
| 2 | 0 | (0.00%) | 2 | (1.02%) | 1 | (0.46%) | 3 |
| 3 | 0 | (0.00%) | 1 | (0.51%) | 0 | (0.00%) | 1 |

When holding other factors as constants, the odds ratio of using first-person pronouns vs. not using first-person pronouns was 5.80 for tweets in moderate vs. poor data veracity (p<0.001). The ratio increased to 7.89 when we compared good data veracity with poor data veracity (p<0.001). The odds of tweets in moderate and good data veracity that express negative sentiment were 5.59 and 6.33 times, respectively, as that of tweets in poor data veracity (p<0.001 and p<0.001). Similarly, the odds of the presence of drug names and ADRs in the same sentence in tweets in good and moderate data veracity were 6.20 and 3.77 times, respectively, as that of tweets in poor data veracity (p<0.001 and p=0.002). No significant difference was observed when we evaluated the effect of using second-person pronouns, number of drugs, and number of indications in tweets between different data veracity levels. However, the differences in the number of drugs and indications became significant at 0.1 and 0.05 significance level when we only compared tweets in poor and good data veracity (p=0.053 and p=0.047). The odds ratio of tweets with good data veracity was multiplied by 0.50 for every one more drug mentioned in the tweets (p=0.053), and multiplied by 0.34 for every one more indication mentioned in the tweets (p=0.047) (Tables 3).

**Table 3.**
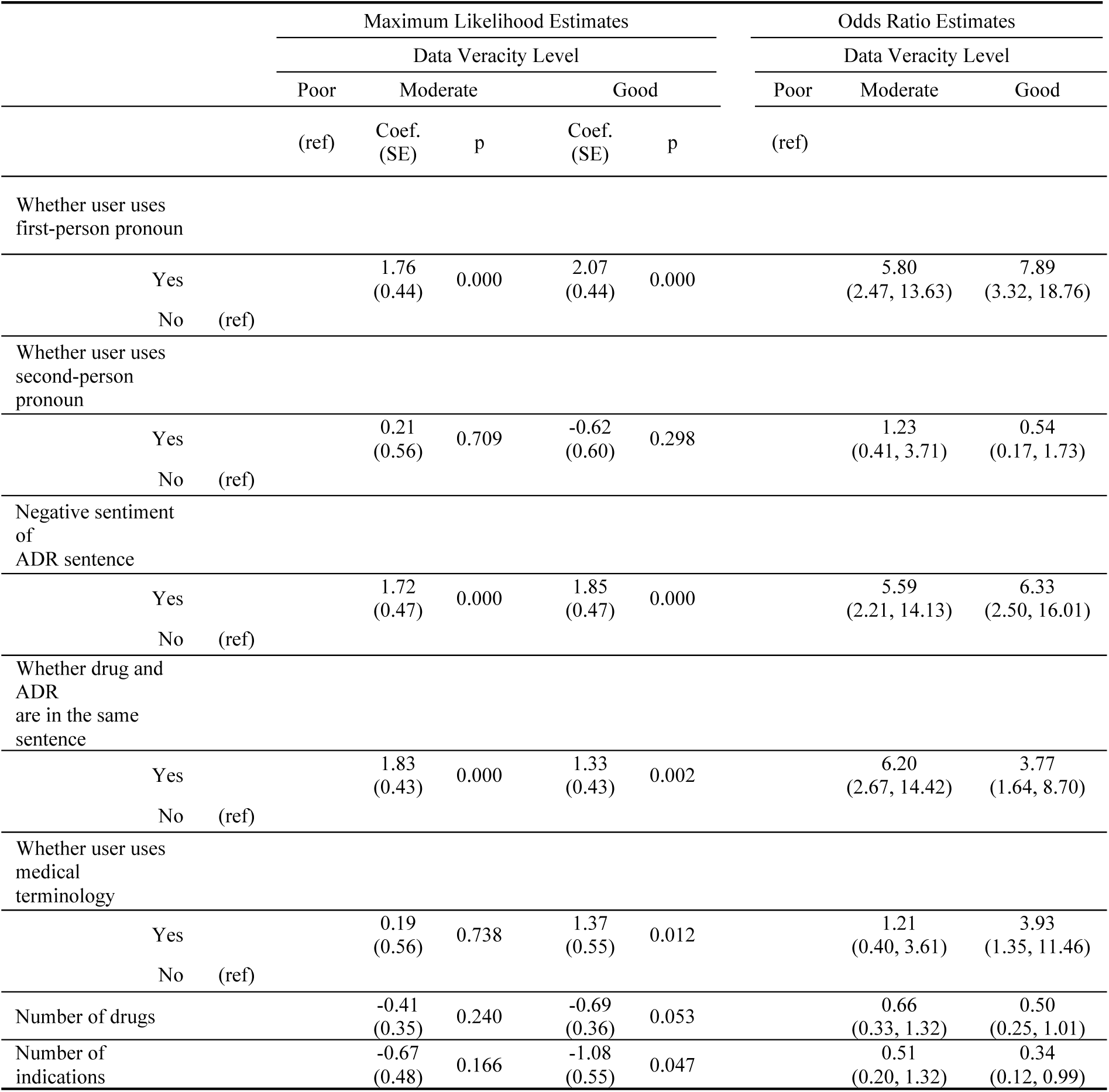
Multinomial Logistic Regression for ADR data veracity levels by tweets characteristics.

## Discussion

In this study, we demonstrated how intricate linguistic features of Twitter posts, when integrated with Twitter variables, can be used to detect diverse data veracity of ADR tweets. Different linguistic features were found to be associated with specific data veracity levels including two that were reported by existing studies: using first-person pronouns; being in negative sentiment. We also found four linguistic features that were not reported elsewhere including the presence of drug and ADRs in the same sentence; using medical terminology; the number of mentioned drugs; and the number of mentioned indications. See Table 4 for a summary. These findings suggest that the quality of Twitter data could be further improved if researchers would use these influential linguistic features to screen ADR-related tweets before used for PV research.

**Table 4.**
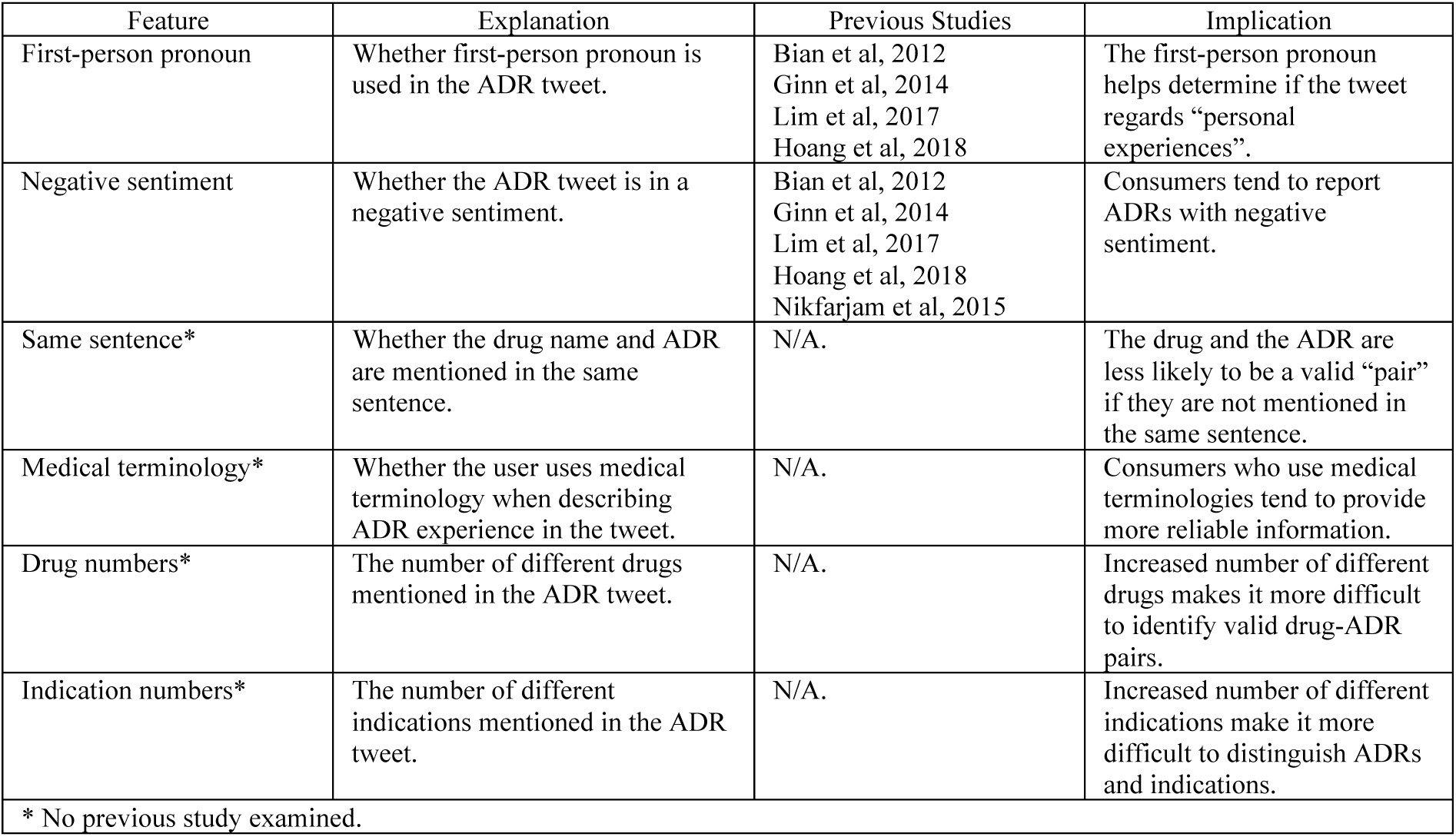
Summary of key linguistic features.

Accurate identification and categorization of key linguistic features in Twitter posts are critical to taking advantages of patients and health consumers reported ADR data. We found several clues of improving data veracity during the progress of annotation. First, drug names or keywords when out of context are causes of poor data veracity. For example, “lozenge” was identified in dataset as a drug, but it was never specified what active ingredients the lozenge contained. When “Nicotine” was identified as a drug with ADRs in tweets, the tweets were most discussing over smoking cessation and not the use of nicotine in a medically prescribed way. Tweets often contain mentions of recreational drugs (e.g., heroin), but such drugs and linked adverse reactions are out of the context of ADR. Second, linguistic features could be mistakenly categorized since the information contained may be incomplete and limited due to the low text limits.^22^ For example, we found instances of a comorbidity being identified as an ADR. Such cases would be more of a challenge since even medical experts might not be able to distinguish between comorbidity and ADR based on limited information. Another situation is that vague descriptions of ADRs (e.g., feeling sick, aching, etc.) cannot be mapped onto drug-ADR pairs in SIDER database. The causes of insufficient information vary and can be better understood if we could tease out the medium level of data veracity further, because tweets with insufficient information tend to be categorized into medium data veracity. Third, idiomatic language and metaphors can also impede the correct recognition of linguistic features, leading to the difficulty in identifying either mentions of ADRs or specific ADRs. For example, “it feels like my brain is melting …” can be a metaphor for drowsiness, dizziness, sluggishness, and other possibilities. To correctly decipher the message in these tweets, analyzing the longitudinal patterns and trends of tweets could be of great help. Fourth, NLP-based sentiment analysis can reveal attitudes and feelings of the users who composed the Twitter posts. However, it lacks accuracy when modeling some special situations. For example, sentiment analysis may generate an opposite result of the truth feeling when consumers use sarcasm in their messages, which also is a cause of low data veracity. Future NLP technique should be focused on the pragmatics of the sentence instead of semantics solely. Specifically, studies should pay specific attention to the logical conjunctions and take into account of temporal patterns and trends of semantics across tweets posted by the same users and generated in different times.

These identified linguistic features, Twitter variables, and association rules are key to identifying different data veracity, yet screening Twitter data in the real-world is costly considering that Twitter data consist of more than 50 variables including the Twitter posts in free text and the high volume of dataset needed for PV research. To bridge this gap, our findings suggest great potentials of developing an efficient machine learning model to automatically detect ADR tweets at different levels of data veracity. This model can greatly reduce the human labor when used for Twitter data selection. Our findings shed light on a couple of key designing components for developing machine learning models. First, the numbers of annotated drugs and indications could be used in the machine learning models as our statistical results showed that they are strong predictors for good data veracity tweets. Second, tweets with first person pronoun, negative sentiment of ADR sentence, drug name and ADR name in the same sentence, mentions of drug name, and no mention of indication tend to be of good data veracity. Thus, these linguistic features should be included in the machine learning models as well. Third, annotation of words and phrases using terminologies tailored for layperson’ language (e.g., Consumer Health Vocabulary) could be a strict filter for tweets with low data veracity. Precise distinguishing between indications and ADRs is also important for machine learning models but is not fully addressed in this study.

While the results demonstrated the pathway to data veracity improvement of ADRs from Twitter, our study is subject to limitations. The dataset we used does not contain longitudinal ADR tweets. Temporal dynamics of ADR tweets can be valuable information for assessing the data veracity, which will be addressed in our future study. Additionally, we were unable to assess whether the user-deleted ADR tweets would have any systemic impact on data veracity. Follow-up studies should employ multi-source datasets for cross-validation. Despite limitations, our study is among the first that incorporated the rule-based logic flow generated by medical experts in defining and identifying data veracity levels of ADR tweets, which has the potential of being generalized to other platforms of social media data.

## Data Availability

NA

## Acknowledgements

This study is supported by a seed grant by University of South Carolina Arnold School of Public Health.

